# Research Letter: Characterizing spin in psychiatric clinical research literature using large language models

**DOI:** 10.1101/2024.06.30.24309737

**Authors:** Roy H. Perlis

**Author notes:** **Correspondence:** Roy H. Perlis, Massachusetts General Hospital 185 Cambridge Street, 6th Floor Boston, MA 02114.

## Abstract

**Importance:** Spin is a common form of biased reporting that misrepresents study results in publications as more positive than an objective assessment would indicate, but its prevalence in psychiatric journals is unknown.

**Objective:** To apply a large language model to characterize the extent to which original reports of pharmacologic and non-pharmacologic interventions in psychiatric journals reflect spin.

**Design:** We identified abstracts from studies published between 2013 and 2023 in 3 high-impact psychiatric journals describing randomized trials or meta-analyses of interventions.

**Main Outcome and Measure:** Presence or absence of spin estimated by a large language model (GPT4-turbo, turbo-2024-04-09), validated using gold standard abstracts with and without spin.

**Results:** Among a total of 663 abstracts, 296 (44.6%) exhibited possible or probable spin – 230/529 (43.5%) randomized trials, 66/134 (49.3%) meta-analyses; 148/310 (47.7%) for medication, 107/238 (45.0%) for psychotherapy, and 41/115 (35.7%) for other interventions. In a multivariable logistic regression model, reports of randomized trials, and non-pharmacologic/non-psychotherapy interventions, were less likely to exhibit spin, as were more recent publications

**Conclusions and Relevance:** A substantial subset of psychiatric intervention abstracts in high-impact journals may contain results presented in a potentially misleading way, with the potential to impact clinical practice. The success in automating spin detection via large language models may facilitate identification and revision to minimize spin in future publications.

## Introduction

Spin has been shown to be common in medical publications^1^, a form of biased reporting which misrepresents study results as more positive than an objective assessment would indicate^2^. Despite the attention this tactic has received, a recent study suggested rates of spin remain high^3^.

Because abstracts may be the primary means by which many clinicians interact with the medical literature^4,5^, the presence of spin in abstracts may be particularly consequential: Readers of abstracts with spin are more likely to believe a study yielded positive results^5^. Abstracts with overly positive presentation of results risk distortion of prescribing practices or adoption of new technologies that may not be warranted by the evidence.

As spin has received little attention in the psychiatric literature, we developed an automated method using large language models to detect spin in abstracts related to psychiatric treatment.

## Methods

We used the Biopython Entrez package to identify original research in Pubmed reflecting clinical trials and meta-analyses related to interventions between 2013 and 2023 in the 3 highest-impact psychiatric journals publishing trials: *American Journal of Psychiatry, Lancet Psychiatry*, and *JAMA Psychiatry* (Supplemental Methods).

We applied a large language model (GPT4-turbo, or turbo-2024-04-09) with temperature set at 0, via Python script accessing the chat.ai application programming interface (API). The prompt included a definition of spin^2^ a typology of spin^1^(Supplemental Methods), with a request to characterize the abstracts in terms of presence of spin.

We validated the prompt by presenting 30 gold-standard abstracts characterized as representing spin, and 30 edited to eliminate spin^5^, 4 times each in random order. We then presented each of the psychiatry journal abstracts.

We used logistic regression to examine whether journal, study type, intervention type, and year was significantly associated with likelihood of including spin. Analyses used R 4.3.2. The study did not constitute human subjects research.

## Results

To validate the spin-detection prompt, among the 60 gold standard abstracts, sensitivity was 100% (95% CI, 98-100%) and specificity 91% (95% CI 86-95%); overall accuracy was 230/240 (95.8%).

Among a total of 663 abstracts, 296 (44.6%) exhibited possible or probable spin – 230/529 (43.5%) randomized trials, 66/134 (49.3%) meta-analyses; 148/310 (47.7%) for medication, 107/238 (45.0%) for psychotherapy, and 41/115 (35.7%) for other interventions. In a multivariable logistic regression model, reports of randomized trials, and non-pharmacologic/non-psychotherapy interventions, were less likely to exhibit spin, as were more recent publications (Figure 1).

**Figure 1.**
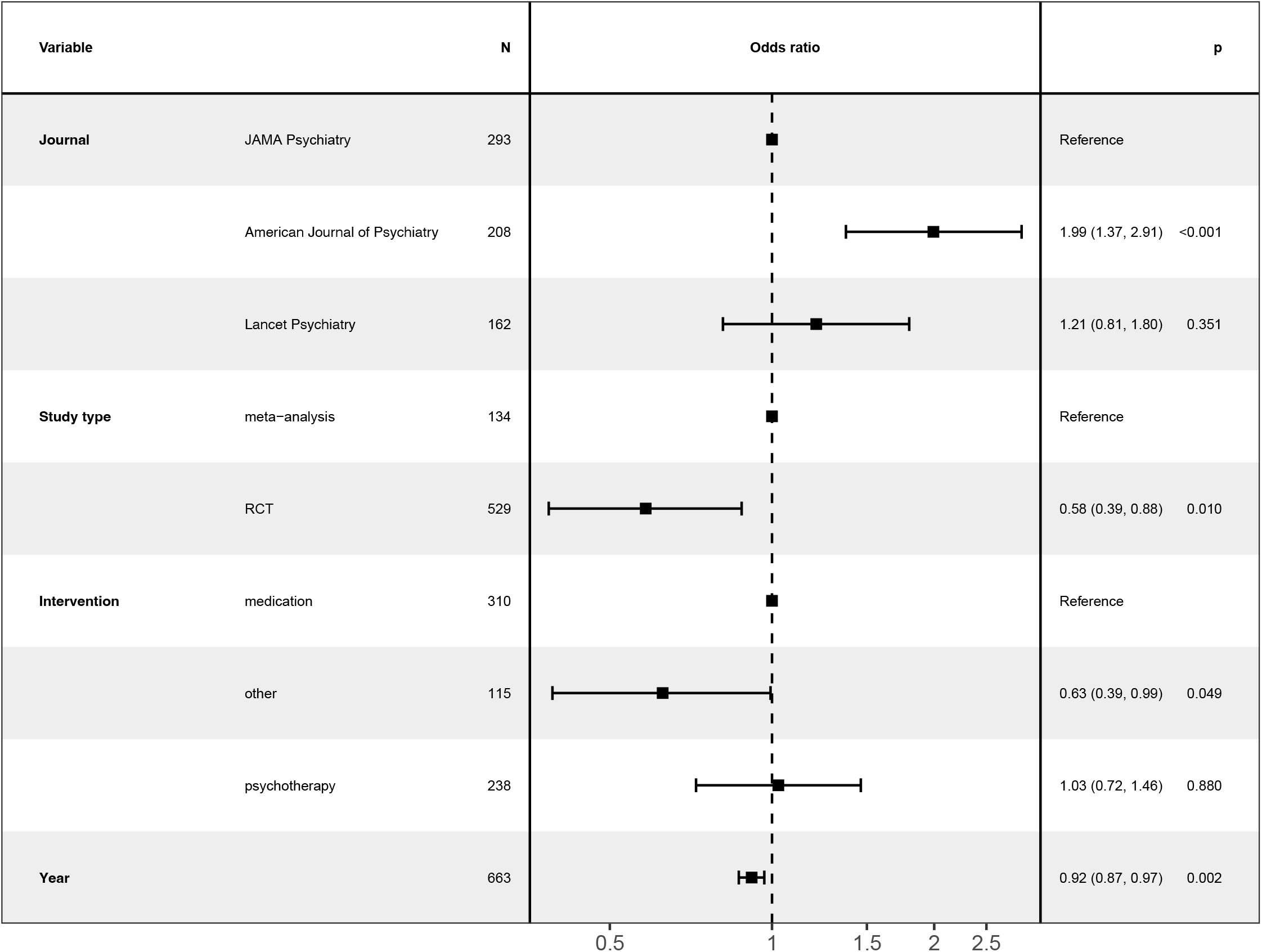
Logistic regression model of abstracts containing possible spin

## Discussion

In this application of a large language model to characterize spin in psychiatric abstracts, we found such language to be relatively common, particularly in meta-analyses, but diminishing over time. One recent study of randomized trials in endometriosis found rates of spin in abstracts to be increasing over the past decade^3^, while an investigation of systematic reviews in melanoma found modest diminution over time, although overall ∼40% of abstracts reviewed included such language^6^. We were unable to identify a prior report in psychiatry.

Our work has multiple limitations. While we demonstrate and validate a novel application of large language models, the imperfect specificity may lead us to overestimate spin. Our sampling frame is restricted to a subset of psychiatry journals; it is possible that spin is more prevalent in journals that publish clinical studies less frequently.

Nonetheless, this investigation suggests that a substantial subset of psychiatric intervention abstracts in high-impact journals may contain results presented in a potentially misleading way, with the potential to impact clinical practice. The success in automating spin detection via large language models may facilitate identification and revision to minimize spin in future publications.

## Data Availability

Data will be available upon reasonable request to the author after publication

## Acknowledgements

Dr. Perlis is supported in part by the National Institute of Mental Health (R01MH123804, U01MH136059). The sponsors did not contribute to the design and conduct of the study; collection, management, analysis, and interpretation of the data; preparation, review, or approval of the manuscript; and decision to submit the manuscript for publication. Dr. Perlis had full access to all the data in the study and takes responsibility for the integrity of the data and the accuracy of the data analysis.

## References

1. Yavchitz A, Ravaud P, Altman DG, et al. A new classification of spin in systematic reviews and meta-analyses was developed and ranked according to the severity. J Clin Epidemiol. 2016;75:56–65. doi:10.1016/j.jclinepi.2016.01.020

2. Latronico N, Metelli M, Turin M, Piva S, Rasulo FA, Minelli C. Quality of reporting of randomized controlled trials published in Intensive Care Medicine from 2001 to 2010. Intensive Care Med. 2013;39(8):1386–1395. doi:10.1007/s00134-013-2947-3

3. Shirafkan H, Moher D, Mirabi P. The reporting quality and spin of randomized controlled trials of endometriosis pain: Methodological study based on CONSORT extension on abstracts. PLOS ONE. 2024;19(5). doi:10.1371/journal.pone.0302108

4. Barry HC, Ebell MH, Shaughnessy AF, Slawson DC, Nietzke F. Family physicians’ use of medical abstracts to guide decision making: style or substance? J Am Board Fam Pract. 2001;14(6):437–442.

5. Boutron I, Altman DG, Hopewell S, Vera-Badillo F, Tannock I, Ravaud P. Impact of Spin in the Abstracts of Articles Reporting Results of Randomized Controlled Trials in the Field of Cancer: The SPIIN Randomized Controlled Trial. JCO. 2014;32(36):4120–4126. doi:10.1200/JCO.2014.56.7503

6. Nowlin R, Wirtz A, Wenger D, et al. Spin in Abstracts of Systematic Reviews and Meta-analyses of Melanoma Therapies: Cross-sectional Analysis. JMIR Dermatology. 2022;5(1):e33996. doi:10.2196/33996

