## Supplemental Materials for "Research Letter: Characterizing spin in psychiatric clinical research literature using large language models"

Supplemental Methods

**Pubmed query:**

(("JAMA Psychiatry"[Journal]) OR ("Am J Psychiatry"[Journal]) OR ("Archives of General Psychiatry"[Journal]) OR ("Lancet Psychiatry"[Journal])) AND (("clinical trial"[Publication Type]) OR ("meta-analysis"[Publication Type])) AND (2013/01/01:2023/12/31[PDAT]) AND (("treatment"[Title/Abstract]) OR ("medication"[Title/Abstract]) OR ("complementary medicine"[Title/Abstract]) OR ("ECT"[Title/Abstract]) OR ("TMS"[Title/Abstract]) OR ("ketamine"[Title/Abstract]) OR ("psychotherapy"[Title/Abstract]) OR ("intervention"[Title/Abstract]))

**LLM prompt (incorporates text from references 1 and 5; see main text for details):**

Spin is defined as a way of reporting to convince readers that a treatment was beneficial, despite a statistically nonsignificant difference. Types of spin:

{Yavchitz et al (reference #1), Table 1, right column}

Statistically significant results should not be labeled as spin for categories focusing on reporting significance. A wide confidence interval is defined as crossing the null value by a substantial margin. A small effect size is defined as a clinically insignificant difference. Consider the context and full content of the abstract when identifying spin. Note that conclusions stating no significant difference in outcomes should not be labeled as spin for categories 6, 12, and 14. Specifically:

- Sentences that indicate no statistically significant difference between treatment groups should not be considered spin for categories 6, 12, and 14.

- Sentences that provide statistically significant results with clear confidence intervals and p-values should not be considered spin for any category.

- Sentences that clearly and accurately report nonstatistically significant results should not be labeled as spin.

- Sentences that report statistically significant results with p-values and confidence intervals should not be considered spin, especially for categories 6 and 12.

- Do not assign spin if #6 and/or #12 are the only concerning categories.

Now, for the following abstract, please only return a JSON object with 4 key-value pairs, with no other text: study design (RCT, naturalistic, meta-analysis); treatment studied (medication, psychotherapy, other); likelihood of spin (no, possible, probable) where possible is less than 50% confidence; sentence or phrase in abstract most worrisome for spin, if any; categories of spin in list form (#1-21 from the prompt). Example output: {"design": "RCT", "treatment":"medication","spin":"probable","sentence":"key sentence","categories":"[6,15]"} ...

**Illustration of interactive application:**

*User:* why is this spin?

*ChatGPT (GPT4o):*

The determination of spin in this abstract is based on the presence of several types of spin as defined in the prompt. Here's a breakdown of why it was labeled as "probable" spin:

1. **Selective reporting of or overemphasis on efficacy outcomes favoring the beneficial effect of the experimental intervention (#1)**:
   - The abstract emphasizes that [treatment] demonstrated antidepressant efficacy, highlighting the significantly greater improvement in HAM-D and HAM-A scores, while downplaying the fact that there was no significant improvement in the Visual Analog Scale for Pain score.
2. **Conclusion focusing selectively on statistically significant efficacy outcome (#6)**:
   - The conclusion claims efficacy based on statistically significant results for some measures (HAM-D and HAM-A), but does not adequately address the non-significant results for the Visual Analog Scale for Pain score or the 14% discontinuation rate due to intolerance in the [treatment] group.
3. **Conclusion claiming the beneficial effect of the experimental treatment despite high risk of bias in primary studies (#15)**:
   - The conclusion states that [treatment] demonstrated antidepressant efficacy without discussing the potential biases, such as the 14% discontinuation rate due to intolerance, which could impact the overall assessment of the treatment's efficacy and safety.

These factors contribute to the likelihood of spin, as they present the treatment in a more favorable light than might be warranted by the complete set of results.

*User:* Rewrite the conclusion to not exhibit spin

*ChatGPT:*

Certainly. Here's a revised conclusion that aims to present a balanced and comprehensive view of the study's findings:

**Original Conclusion:** "[Treatment] as an adjunct to escitalopram demonstrated antidepressant efficacy in adult patients with major depressive disorder experiencing persistent symptoms after 8 weeks of open-label treatment with escitalopram."

**Revised Conclusion:** "Adjunctive [treatment] to escitalopram showed significant improvement in depression severity as measured by the HAM-D and HAM-A scores in adult patients with major depressive disorder who had persistent symptoms after 8 weeks of open-label treatment with escitalopram. However, there was no significant improvement in pain scores, and a notable percentage (14%) of patients in the [treatment] group discontinued treatment due to intolerance. These findings suggest potential benefits of adjunctive [treatment], but also highlight the need for careful consideration of its tolerability and the mixed results across different outcome measures."

**References**

1.	Yavchitz A, Ravaud P, Altman DG, et al. A new classification of spin in systematic reviews and meta-analyses was developed and ranked according to the severity. *J Clin Epidemiol*. 2016;75:56-65. doi:10.1016/j.jclinepi.2016.01.020

5.	Boutron I, Altman DG, Hopewell S, Vera-Badillo F, Tannock I, Ravaud P. Impact of Spin in the Abstracts of Articles Reporting Results of Randomized Controlled Trials in the Field of Cancer: The SPIIN Randomized Controlled Trial. *JCO*. 2014;32(36):4120-4126. doi:10.1200/JCO.2014.56.7503
